# Prospective evaluation of basic life support interventions for foreign body airway obstructions: A pilot population-based cohort study

**DOI:** 10.64898/2026.09.18.26363413

**Authors:** Cody L Dunne, Jenice Tea, Beatrice Rivera, Ian E Blanchard, Catherine Patocka, Khara Sauro, Andrew D McRae

**Author notes:** Co-senior author.

## Abstract

**Background:** Foreign body airway obstruction (FBAO) is a life-threatening emergency; however, bystander treatment recommendations rely on retrospective data. We evaluated the feasibility of a population-based prospective cohort study of FBAO interventions.

**Methods:** We conducted a prospective observational pilot study in Alberta, Canada, between November 2025 and April 2026. We identified individuals with out-of-hospital FBAO who called emergency medical services or attended an emergency department using a validated health-record case definition. Participants were stratified into adults and children (aged>2 years) and younger children (aged≤ 2 years) cohorts. The primary outcome was included cases per month, with several secondary feasibility (proportion of eligible cases included, data completeness) and clinical outcomes (FBAO relief, survival to acute care discharge, favourable neurological outcome, intervention-associated injuries).

**Results:** Among 5,208 screened encounters, 147 FBAO cases were identified. Of these, 91 adults and children (89.2%) and 39 young children (86.7%) were enrolled. Mean recruitment was 15.2 participants/month (SD=1.1) in the adult and child cohort, exceeding the predefined feasibility threshold (≥13/month), and 6.5 participants/month (SD=1.3) in the young child cohort, meeting the amber-zone criterion (6-8/month). Data completeness exceeded 90% across all participants. In adults and children, bystander intervention relieved the obstruction in 59 cases (66.3%), and 80(87.9%) survived to acute care discharge. In young children, bystander intervention relieved the obstruction in 33 cases (84.6%); all survived with favourable neurological outcomes.

**Conclusions:** Prospectively identifying and recruiting individuals with FBAO is feasible using population-based prehospital and emergency department records. These findings support a larger adequately powered prospective cohort study comparing the effectiveness and safety of FBAO interventions.

## Introduction

Foreign body airway obstructions (FBAO, choking) are life-threatening medical emergencies, which will result in death rapidly if not treated (1–6). Bystander intervention significantly improves survival, as mortality rises sharply when the FBAO is not removed before paramedic arrival (1,6). Despite this, there exists considerable uncertainty over which bystander intervention is most effective. Current treatment recommendations for FBAO differ based on age category and resuscitation organization (7–11); however, all include some combination of back blows, abdominal thrusts, and/or chest thrusts.

The historically limited data in this field underlies the inconsistent treatment recommendations and may lead to uncertainty when treating patients (12). Until recently, treatment recommendations were based on case series and no study compared the relative effectiveness or safety of different FBAO BLS interventions (12). In 2024, a Canadian study compared FBAO interventions using retrospective data and found back blows improved FBAO relief and survival to discharge (6). However, retrospective data may incompletely capture the timing, sequence, and outcomes of FBAO interventions. In 2025, a Japanese study reported the first prospective collection of FBAO intervention data, however their work included only cases transported to emergency departments and did not compare specific FBAO interventions (13).

There is an urgent need for prospective data comparing the effectiveness and safety of FBAO interventions across both prehospital and emergency department settings to inform treatment recommendations. However, the feasibility of such a study has not been evaluated to date. Accordingly, we conducted a pilot prospective cohort study to determine the feasibility of prospective FBAO case identification and recruitment while exploring preliminary intervention and outcome data.

## Methods

### Study Design and Setting

This observational pilot study used electronic health records and patient interviews to assess the feasibility of conducting a prospective cohort study on FBAO basic life support (BLS) interventions. This study is reported according to the Strengthening the Reporting of Observational Studies in Epidemiology (STROBE) guidelines and the relevant sections of the Consolidated Standards of Reporting Trials (CONSORT) extension to randomised pilot and feasibility trials guidelines (14,15). The Conjoint Health Research Ethics Board of the University of Calgary (REB25-0419) approved the study. The study was registered a priori on ClinicalTrials.gov (NCT07365137 and NCT07348848) (16,17).

The study used population-level data from Alberta, Canada, between November 1, 2025, and April 30, 2026. In Alberta, during the study period, Alberta Health Services was the only health services payer and coordinated all publicly-funded health services, including emergency medical services (EMS), to all residents of the province. At birth (if born in Alberta) or when they register as a resident (if relocated to the province), the province assigns a unique healthcare number to each individual allowing direct linkage of all healthcare encounters.

### Cohort Identification and Data Sources

We included individuals of all ages who experienced an out-of-hospital FBAO and called emergency medical services (EMS) or attended an emergency department during the study period. We excluded individuals with abnormal airway anatomy, such as a tracheostomy.

We defined FBAO cases as individuals who presented with a history of an object or substance not native to the airway introduced, followed by clinical signs of obstruction (e.g., stridor, cyanosis, hypoxemia, inability to move air).

FBAO cases were identified using the EMS electronic Patient Care Record (ePCR) (Siren ePCR Version 4, Medusa Medical Technologies, Halifax, NS) and the Alberta Health Services’ Connect Care clinical information system (Connect Care Clinical Information System, Alberta Health Services, Edmonton, AB). The ePCR is the single electronic medical record system for all prehospital providers in Alberta and includes demographics, vital signs, and standardized reporting of incident, patient, and treatment details. It also includes a free-text narrative synopsis, including information on the events preceding the FBAO, interventions performed by bystanders, patient assessment conducted by the paramedic, and all subsequent interventions. Connect Care is the single acute electronic medical record system for all emergency department visits and hospitalizations in Alberta. Both include population-based data for the province.

We identified potential FBAO cases from the ePCR and Connect Care using a previously validated algorithm, employing a combination of the paramedic’s primary impression, treatment protocol, interventions provided, and emergency department chief complaint (5,6). An emergency medicine physician (CD) subsequently reviewed all available case data to determine true FBAO cases.

Once FBAO cases were identified, the individual (or their parent or guardian for pediatric cases) were contacted by a healthcare provider to invite them to participate in a virtual interview. If agreeable, a research assistant followed up to obtain informed consent and complete an interview on their FBAO incident. Research assistants used a standardized data collection tool for each interview. We pilot tested the data collection tool among five individuals without healthcare or research training to ensure laypersons easily understood each question before recruitment started. In the case that an individual was unable to participate in an interview due to severe injury, death, or inability to be contacted, then the Research Ethics Board granted a waiver of consent to use the individual’s health records for data collection.

Given differences in age-specific FBAO treatment recommendations (7,10), we created two cohorts for the purpose of the pilot study: adults and children (aged older than 2 years) and younger children (aged 2 years or younger). Children aged 1-2 years were grouped with the young child cohort because previous Alberta data demonstrated they frequently receive interventions consistent with infant treatment recommendations (6).

### Variables

Our primary outcome was the included case rate per month. Our secondary feasibility outcomes were the proportion of eligible cases included, and the proportion of cases with greater than 90% of variables collected. Our secondary exploratory clinical outcomes were FBAO relief, survival to acute care discharge (i.e., EMS, emergency department, or hospital discharge), survival to acute care discharge with favourable neurological outcome, and intervention-associated injuries.

We defined FBAO relief as present if the responders determined that the case did not require further FBAO intervention due to improvement or resolution of their respiratory distress. For individuals in cardiorespiratory arrest, we determined FBAO relief if responders were able to ventilate successfully after an intervention where previously they could not. This was adjudicated from available health records and interview data. We initially defined survival to discharge with favourable neurological outcome in the protocol based on individual activities of daily living status (16,17), however, prior to data collection we decided instead to use a global assessment based on the adult and pediatric cerebral performance categories (CPC) and defined favourable neurological outcome as no change in CPC between pre- and post-FBAO incident (18,19).

For the clinical secondary outcomes, our exposure variable was FBAO intervention coded as abdominal thrusts, back blows, chest thrusts (when performed on a conscious person), cardiopulmonary resuscitation (CPR) (when chest compressions were performed on an unconscious person), or a combination of these. Appendix A defines all other variables collected and possible values.

### Sample Size

#### Adult and Child Cohort

Based on our prior work (6), using previously observed success proportions (abdominal thrusts: 59.5% versus back blows: 72.2%) and expecting abdominal thrusts to be used about 3.5 times more often than back blows, we estimated that 640 cases would be needed to obtain 80% power, with an alpha of 0.05 using the two-sided z-test of independent proportions. We used abdominal thrusts and back blows for the calculation given most adult and child FBAO cases in Alberta receive only one of these interventions (6).

We deemed 48 months an acceptable period to complete data collection given the relatively low frequency of FBAO incidents in our prior work (6). Therefore, based on our sample size calculation, we would need to include a mean of 13.3 cases per month (640 cases divided by 48 months). We deemed the slowest acceptable period would be one additional year of data collection (60 months total), which would equate to a mean of 10.6 cases per month.

Using the traffic light approach for a feasibility study’s sample size, we set the green criterion (progress to full study, no concerns) to 13 cases or more per month and the red criterion (major problems, potentially not remediable concerns) to less than 10 cases per month. The amber zone was set as 10-13 cases per month, which represented a recruitment rate range that may be compatible with a successful future study; however, we would first aim to identify and address remediable recruitment challenges before deciding whether to proceed (20).

#### Young Child Cohort

Using a similar approach, we estimated that 381 cases would be needed to obtain 80% power, with an alpha of 0.05 using the two-sided z-test of independent proportions to compare back blows and chest thrusts with previously observed success of 76.5% and 55.6%, respectively, with a ratio of use equal to 8.2 (6). Back blows and chest thrusts were selected for comparison given they are the most frequently used interventions for young children in Alberta (6). This resulted in a green criterion of 8 cases per month, a red criterion of 6 cases included per month, and an amber zone of 6-8 cases per month.

### Analysis

We summarized patient and situation characteristics using descriptive statistics, with frequencies (proportions) or medians (interquartile range [IQR]) as appropriate.

Our primary feasibility outcome (included case rate) was calculated by obtaining monthly included cases counts and summarized as mean (standard deviation [SD]) over the six-month study period. Our secondary feasibility outcomes (eligible cases included and number of cases with greater than 90% of variables collected) are reported as proportions of the total number of cases identified and included, respectively.

As the pilot study was underpowered to accurately estimate the precision of our clinical outcomes, we reported these using only descriptive statistics (frequencies with proportions) stratified by bystander intervention.

We presented all data and outcomes separately for each age cohort. All analyses were performed using STATA 16 (StataCorp. 2021. Stata Statistical Software: Release 16. College Station, TX: StataCorp LLC).

## Results

During the study period, 5208 encounters were screened and 147 were identified as true FBAO cases (2.9%) (Figure 1). Of these, there were 102 (69.4%) individuals older than two years (adult and child cohort) and 45 (30.6%) individuals aged two years or younger (young child cohort). After enrollment, 11 and 6 individuals (or their parent/guardian) declined to participate in the adult and child or young child cohort, respectively. As a result, we included 91 individuals in the adult and child cohort (89.2%) and 39 individuals in the young child cohort (86.7%).

**Figure 1:**
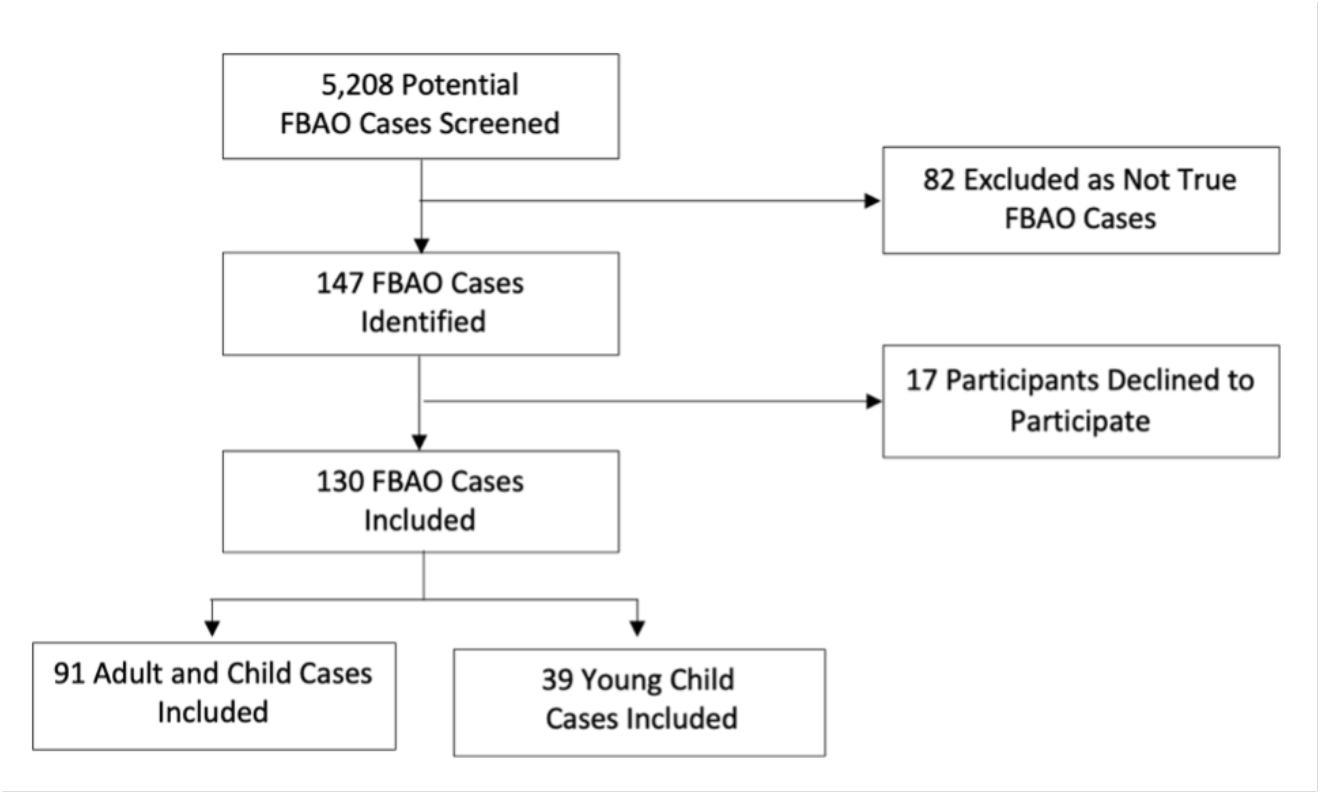
Patient flow diagram. *FBAO = Foreign body airway obstruction*

### Adult and Child Cohort

We included a mean of 15.2 individuals per month (standard deviation [SD] = 1.1), and 89.2% of eligible individuals during the study period. All included individuals had greater than 90% of pre-determined variables recorded, with the most frequently missing variables relating to the pre-EMS bystander (e.g., age, height, weight) (Appendix A, Table A1).

The adult and child cohort was a median age of 71 years (IQR 55-84), with slight female predominance (n=52, 57.1%) (Table 1). Approximately one-third of individuals had previously had a FBAO (n=31, 34.1%), while a diagnosis of major neurocognitive disorder (n=31, 34.1%) or dysphagia (n=29, 31.9%) were common. Thirty-seven (40.7%) individuals were care facility residents, with 54 (60.0%) and 79 (87.8%) individuals independent with mobility and eating, respectively.

**Table 1:** Patient, situation, and intervention characteristics for each FBAO prospective cohort. CPR = Cardiopulmonary resuscitation; EMS = Emergency medical services; FBAO = Foreign body airway obstruction; IQR = Interquartile range; LOC = Level of consciousness; N/a = Not applicable ^1^Independent as appropriate for age ^2^Interventions applied in any order

| Characteristic | Adult and Child Cohort (n [%])<br>N = 91 | Young Child Cohort (n [%])<br>N = 39 |
| --- | --- | --- |
| **Median Age (IQR)** |  |  |
|  | 71 years (55-84) | 9 months (2-15) |
| **Age Category** |  |  |
| Younger than 1 year | N/a | 24 (61.5) |
| 1 – 2 years | N/a | 15 (38.5) |
| 3 – 12 years | 13 (14.3) | 0 (0) |
| 13 – 17 years | 0 (0) | 0 (0) |
| 18 – 64 years | 21 (23.1) | 0 (0) |
| 65 years or older | 57 (62.6) | 0 (0) |
| **Female Sex** |  |  |
|  | 52 (57.1) | 11 (28.2) |
| **Median Patient Weight, kg (IQR)** |  |  |
|  | 68.5 (53-80) | 9 (5-11) |
| **Median Patient Height, cm (IQR)** |  |  |
|  | 162 (155-175) | 66.5 (55-73) |
| **Medical Comorbidities** |  |  |
| Prior FBAO | 31 (34.1) | 2 (5.1) |
| Major neurocognitive disorder | 31 (34.1) | 0 (0) |
| Dysphagia | 29 (31.9) | 0 (0) |
| Movement disorder | 10 (11) | 0 (0) |
| Stroke | 12 (13.2) | 0 (0) |
| Median Charlson Comorbidity Index (IQR) | 4 (2-6) | 0 (0-0) |
| **Activities of Daily Living** |  |  |
| Independent with eating | 79 (87.8) | 39 (100)<sup>1</sup> |
| Independent with mobility | 54 (60) | 39 (100)<sup>1</sup> |
| **Care facility resident** |  |  |
|  | 37 (40.7) | 0 (0) |
| **Foreign body type** |  |  |
| Liquid food substance | 0 (0) | 16 (41.0) |
| Non-food object | 8 (8.9) | 6 (15.4) |
| Solid food substance | 82 (91.1) | 17 (43.6) |
| **Geographical Location** |  |  |
| Home | 45 (49.5) | 36 (92.3) |
| Long-term care facility or equivalent | 37 (40.7) | 0 (0) |
| Restaurant | 4 (4.4) | 0 (0) |
| School, work, or daycare | 1 (1.1) | 2 (5.1) |
| Other public place | 4 (4.4) | 1 (2.6) |
| **Witnessed FBAO** |  |  |
|  | 82 (90.1) | 36 (92.3) |
| **Pre-EMS Bystander Response** |  |  |
|  | 89 (97.8) | 39 (100) |
| **Bystander** |  |  |
| Family or caregiver | 43 (48.3) | 37 (94.9) |
| Lay bystander | 9 (10.1) | 2 (5.1) |
| Self | 1 (1.1) | 0 (0) |
| Trained bystander with duty to respond | 36 (40.5) | 0 (0) |
| **Bystander Training** |  |  |
| No training | 1 (2.4) | 1 (7.1) |
| First aid or CPR | 5 (11.9) | 10 (71.4) |
| Advanced Training | 36 (85.7) | 3 (21.4) |
| **Patient's Initial Status with Bystander** |  |  |
| Severe FBAO | 81 (91.0) | 37 (94.9) |
| Decreased LOC or unconscious | 8 (9.0) | 2 (5.1) |
| **Initial Bystander Intervention** |  |  |
| Abdominal thrusts | 68 (76.4) | 1 (2.6) |
| Back blows | 12 (13.5) | 37 (94.9) |
| Chest thrusts or compressions on conscious individual | 2 (2.3) | 0 (0) |
| CPR on unconscious individual | 7 (7.9) | 0 (0) |
| LifeVac airway clearance device | 0 (0) | 1 (2.6) |
| **All Bystander Interventions Applied<sup>2</sup>** |  |  |
| Abdominal thrusts | 49 (55.1) | 0 (0) |
| Back blows | 11 (12.4) | 33 (84.6) |
| Chest thrusts on conscious individual | 2 (2.3) | 0 (0) |
| CPR on unconscious individual | 7 (7.9) | 0 (0) |
| LifeVac airway clearance device | 0 (0) | 1 (2.6) |
| Abdominal thrusts and back blows | 10 (11.2) | 0 (0) |
| Abdominal thrusts and chest thrusts | 0 (0) | 1 (2.6) |
| Abdominal thrusts and CPR | 9 (10.1) | 0 (0) |
| Abdominal thrusts and LifeVac | 1 (1.1) | 0 (0) |
| Back blows and chest thrusts | 0 (0) | 4 (10.3) |
| **EMS Called** |  |  |
|  | 84 (92.3) | 29 (74.4) |
| **Ongoing FBAO upon EMS Arrival** |  |  |
|  | 15 (16.5) | 2 (6.9) |
| **Patient's Status upon EMS Arrival** |  |  |
| Severe FBAO | 2 (13.3) | 1 (50) |
| Decreased LOC or unconscious | 4 (26.7) | 1 (50) |
| Cardiac arrest | 9 (60) | 0 (0) |
| **EMS Interventions** |  |  |
| Any intervention | 14 (93.3) | 2 (100) |
| Back blows | 0 (0) | 1 (50) |
| CPR | 8 (57.1) | 0 (0) |
| Laryngoscopy and Magill forceps | 8 (57.1) | 1 (50) |
| Supraglottic airway | 1 (7.1) | 0 (0) |
| Endotracheal intubation | 2 (14.3) | 0 (0) |
| **Prehospital cardiac arrest** |  |  |
|  | 15 (16.5) | 0 (0) |

The majority of FBAO were due to solid food substances (n=82, 91.1%) and occurred in their residence (either home [n=45, 49.5%] or care facility [n=37, 40.7%]). Bystanders witnessed most FBAO (n=82, 90.1%) and responded to them (n=89, 97.8%), frequently before the choking individuals deteriorated to decreased consciousness (n=81, 91%). Bystanders most often initially responded with abdominal thrusts (n=68, 76.4%) or back blows (n=12, 13.5%). When considering all intervention combinations, bystanders used abdominal thrusts only (n=49, 55.1%), back blows only (n=11, 12.4%), and abdominal thrusts combined with back blows (n=10, 11.2%) most frequently.

Few individuals still had a FBAO when paramedics arrived (n=15, 16.5%), with most either unconscious (n=4, 26.7%) or in cardiac arrest (n=9, 60.0%). Paramedics frequently used a combination of CPR (n=8, 57.1%) with laryngoscopy and Magill forceps (n=8, 57.1%) in an effort to relieve the FBAO, however, advanced airway management was seldomly used (n=3, 21.4%) likely due to most of these individuals having advanced care directives excluding airway management (n=10, 66.7%).

#### Bystander Intervention Effectiveness and Safety

The initial intervention relieved the FBAO in 47 individuals (52.8%) (Table 2). Of those who received abdominal thrusts and back blows initially, FBAO relief occurred in 34 (50.0%) and 10 (83.3%) individuals respectively (Table 3). When considering all intervention combinations applied, bystanders relieved the FBAO in 59 (66.3%) individuals, including 33 (67.3%) receiving abdominal thrusts only, 10 (90.9%) receiving back blows only, and 8 (80.0%) receiving abdominal thrusts and back blows.

**Table 2:** Outcomes for each FBAO prospective cohort. EMS = Emergency medical services; ROSC = Return of spontaneous circulation

| Outcome |  | Adult and Child<br>Cohort (n [%])<br>N = 91 | Young Child<br>Cohort (n [%])<br>N = 39 |
| --- | --- | --- | --- |
| **FBAO Relief with Initial Intervention** |  |  |  |
|  |  | 47 (52.8) | 31 (79.5) |
| **FBAO Relief with Multiple Interventions** |  |  |  |
|  |  | 59 (66.3) | 33 (84.6) |
| **EMS Disposition** |  |  |  |
| EMS not called |  | 7 (7.7) | 0 (0) |
| Declined transport |  | 12 (13.2) | 6 (15.4) |
| Resuscitation terminated on scene |  | 11 (12.1) | 0 (0) |
| Transported to hospital |  | 61 (67.0) | 33 (84.6) |
| **Prehospital ROSC** |  |  |  |
|  |  | 4 (26.7) | N/a |
| **Hospital Admission** |  |  |  |
|  |  | 17 (18.7) | 5 (12.8) |
| **Median Hospital Length of Stay, days**<br>**(IQR)** |  |  |  |
|  |  | 6 (4-9) | 6 (1-14) |
| **Intensive Care Admission** |  |  |  |
|  |  | 0 (0) | 2 (5.1) |
| **Survived to Acute Care Discharge** |  |  |  |
|  |  | 80 (87.9) | 39 (100) |
| **Survived to Acute Care Discharge with**<br>**Favourable Neurological Outcome** |  |  |  |
|  |  | 80 (87.9) | 39 (100) |

**Table 3:** FBAO Relief and Survival Outcomes Stratified by Bystander Intervention Received. CPR = Cardiopulmonary resuscitation; FBAO = Foreign body airway obstruction

| Outcome |  | FBAO Relief | Survived to Acute Care Discharge | Survived to Acute Care Discharge with Favourable Neurological Outcome |
| --- | --- | --- | --- | --- |
| **Adult and Child Cohort** |  |  |  |  |
| **Initial Bystander Intervention** |  |  |  |  |
| Abdominal thrusts | N=68 | 34 (50) | 62 (91.2) | 62 (91.2) |
| Back blows | N=12 | 10 (83.3) | 12 (100) | 12 (100) |
| Chest thrusts or compressions on conscious individual | N=2 | 1 (50) | 1 (50) | 1 (50) |
| CPR on unconscious individual | N=7 | 2 (28.6) | 3 (42.9) | 3 (42.9) |
| **All Bystander Interventions** |  |  |  |  |
| Abdominal thrusts | N=49 | 33 (67.3) | 47 (95.9) | 47 (95.9) |
| Back blows | N=11 | 10 (90.9) | 11 (100) | 11 (100) |
| Chest thrusts on conscious individual | N=2 | 1 (50) | 1 (50) | 1 (50) |
| CPR on unconscious patient | N=7 | 2 (28.6) | 3 (42.9) | 3 (42.9) |
| Abdominal thrusts and back blows | N=10 | 8 (80) | 10 (100) | 10 (100) |
| Abdominal thrusts and CPR | N=9 | 4 (44.4) | 5 (55.6) | 5 (55.6) |
| Abdominal thrusts and LifeVac | N=1 | 1 (100) | 1 (100) | 1 (100) |
| **Young Child Cohort** |  |  |  |  |
| **Initial Intervention** |  |  |  |  |
| Abdominal thrusts | N=1 | 0 (0) | 1 (100) | 1 (100) |
| Back blows | N=37 | 30 (81.1) | 37 (100) | 37 (100) |
| LifeVac airway clearance device | N=1 | 1 (100) | 1 (100) | 1 (100) |
| **All Interventions** |  |  |  |  |
| Back blows | N=33 | 29 (87.9) | 33 (100) | 33 (100) |
| LifeVac airway clearance device | N=1 | 1 (100) | 1 (100) | 1 (100) |
| Abdominal thrusts and chest thrusts | N=1 | 0 (0) | 1 (100) | 1 (100) |
| Back blows and chest thrusts | N=4 | 3 (75) | 4 (100) | 4 (100) |

Eighty (87.9%) individuals survived to acute care discharge. All individuals who died had resuscitation terminated on scene, with the majority (n=8, 72.7%) having pre-existing advanced care directives excluding resuscitation. Favourable neurological outcome at acute care discharge occurred in all individuals who survived. Of those who received abdominal thrusts and back blows initially, both survival and favourable neurological outcome occurred in 62 (91.2%) and 12 (100%) individuals, respectively. When considering all intervention combinations, both survival and favourable neurological outcome occurred in 47 (95.9%) individuals receiving abdominal thrusts only, 11 (100%) receiving back blows only, and 10 (100%) receiving abdominal thrusts and back blows.

Intervention-associated injuries were rare (n=4, 4.4%). Two individuals who received abdominal thrusts and CPR, as well as one individual who received abdominal thrusts only, were diagnosed with rib fractures. One individual who received abdominal thrusts and CPR was diagnosed with a hemothorax.

### Young Child Cohort

We included a mean of 6.5 individuals per month (SD = 1.3), and 86.7% of eligible individuals were included during the study period. All included cases had greater than 90% of pre-determined variables recorded, with the most frequently missing variables relating to the pre-EMS bystander (e.g., age, height, weight) (Appendix A, Table A1).

The young child cohort was a median age of 9 months (IQR 2-15), and predominantly male (n=28, 71.8%) (Table 1). Only 2 (5.1%) young children had a prior FBAO. Most FBAO were due to solid food substances (n=17, 43.6%) or liquid food substances (e.g., milk) (n=16, 41.0%), and almost all occurred in their residence (n=36, 92.3%). Bystanders witnessed most FBAO (n=36, 92.3%) and responded to all cases (n=39, 100%).

Bystanders almost always initially responded with back blows (n=37, 94.9%), with one bystander using the LifeVac airway clearance device (2.6%) and one young child receiving abdominal thrusts (2.6%). Only four cases received multiple interventions (n=4, 10.3%).

Few young children still had a FBAO when paramedics arrived (n=2, 6.9%), with none in cardiac arrest. Paramedics used either back blows (n=1, 50%) or laryngoscopy and Magill forceps (n=1, 50.0%) to relieve the FBAO. There were no cases requiring CPR or advanced airway management.

#### Bystander Intervention Effectiveness and Safety

The initial intervention relieved the FBAO in 31 individuals (79.5%) (Table 2). Of those who received back blows initially, FBAO relief occurred in 30 cases (81.1%) (Table 3). When considering all intervention combinations applied, bystanders relieved the FBAO in 33 (84.6%) individuals, including 29 (87.9%) receiving back blows only, three receiving back blows and chest thrusts (75.0%), and one with the use of LifeVac (100%). Abdominal thrusts and chest thrusts did not relieve the FBAO in one case. All individuals survived to acute care discharge with favourable neurological outcome. There were no intervention-associated injuries.

## Discussion

This pilot study demonstrated that prospective identification of FBAO cases and recruitment of these patients across prehospital and emergency department settings is feasible. We successfully identified and included 130 individuals (91 adults and children, and 39 young children) into the cohort. The pilot study exceeded feasibility thresholds in the adult and child cohort and approached feasibility in the young child cohort.

Feasibility for our young child cohort fell within the pre-specified amber zone, suggesting that the study may be feasible but recruitment challenges should be addressed before proceeding (20). Recruitment of patients experiencing an FBAO presents unique challenges because choking incidents are sudden, emotionally distressing, and often involve vulnerable individuals such as young children or frail elderly adults. Further, incidents occur largely prehospital, and do not typically involve follow up with a regular care provider. A scoping review identified that timely follow up, flexible contact methods (e.g., phone vs text messaging vs e-mail), and incentives improve recruitment rates among parents (21). After adopting these strategies and introducing a participation incentive (gift card), our inclusion rate increased from 4.7 participants per month during the first half of the study to 8.3 participants per month during the second half. Our findings are consistent with the broader recruitment literature and suggest that rapid follow-up, flexible communication strategies, and modest incentives may be important considerations when conducting prospective FBAO research, particularly in pediatric populations (21,22).

This pilot study, and the ongoing prospective cohort study that it informed, have the potential to meaningfully influence future FBAO treatment guidelines. Current recommendations continue to rely primarily on retrospective data with substantial risk of bias (1,6,12,23–27). To address methodological limitations in prior research, we have incorporated age-specific cohorts based on prior outcome patterns (6,28,29), distinguished between pre-arrest interventions and arrest management to reduce residual confounding (e.g., chest thrusts/compressions on a conscious person vs CPR), and accounted for baseline neurological status when measuring outcomes (12,13). Further, these research studies will generate the evidence necessary to address several outstanding knowledge gaps in FBAO management, including the relative effectiveness and safety of the initial BLS intervention, whether alternating interventions improves outcomes compared with single-intervention approaches, and the optimal sequence of interventions for achieving survival with favourable neurological outcome. Ultimately, we aim to improve the validity and generalizability of FBAO intervention data to inform future treatment recommendations.

### Limitations

While this study is based on population data, it is limited to one Canadian province and so generalizability to other populations should be interpreted with caution. Additionally, our case identification strategy did not capture individuals with a FBAO who received treatment and did not present to an emergency department or call EMS. These cases represent milder cases with excellent outcomes, and their exclusion will bias intervention effectiveness towards higher likelihood of poor outcomes. However, this study is the first to include individuals with a resolved FBAO that do not get transported with EMS to an emergency department, representing more comprehensive case ascertainment than prior prospective and retrospective studies (13,29).

## Conclusion

In this pilot prospective FBAO intervention study, the adult and child cohort exceeded predefined feasibility thresholds, while the young child cohort reached the amber-zone criterion and demonstrated improved recruitment following protocol modifications. These findings support progression to an adequately powered prospective cohort study capable of comparing the effectiveness and safety of different FBAO interventions to inform future resuscitation guidelines.

## Data Availability

Data is not available for sharing.

## Acknowledgements

We would like to acknowledge Mr. Ryan Lee and the Alberta Health Services EMS Research Committee for assistance with obtaining prehospital data.

